# Ultra-rare *de novo* damaging coding variants are enriched in attention-deficit/hyperactivity disorder and identify risk genes

**DOI:** 10.1101/2023.05.19.23290241

**Authors:** Emily Olfson, Luis C. Farhat, Wenzhong Liu, Lawrence A. Vitulano, Gwyneth Zai, Monicke O. Lima, Justin Parent, Guilherme V. Polanczyk, Carolina Cappi, James L. Kennedy, Thomas V. Fernandez

**Author notes:** Corresponding authors: Emily Olfson and Thomas Fernandez. **Major classification:** biological sciences. **Minor classification:** genetics.

## Abstract

Attention-deficit/hyperactivity disorder (ADHD) is a common and impairing neurodevelopmental disorder in which genetic factors play an important role. DNA sequencing of parent-child trios provides a powerful approach for identifying *de novo* (spontaneous) variants, which has led to the discovery of hundreds of clinically informative risk genes for other neurodevelopmental disorders but has yet to be extensively leveraged in studying ADHD. Here, we conducted whole-exome DNA sequencing in 152 parent-child trios with ADHD and demonstrate for the first time a significant enrichment of rare and ultra-rare *de novo* protein-truncating variants and missense variants predicted to be damaging in ADHD cases compared to unaffected controls. Combining these results with a large independent case-control DNA sequencing cohort (3,206 ADHD cases and 5,002 controls), we identify *lysine demethylase 5B* (*KDM5B)* as a high-confidence risk gene for ADHD as well as two likely risk genes. We estimate that 862 genes contribute to ADHD risk. Finally, using our list of genes harboring ultra-rare *de novo* damaging variants, we show that these genes overlap with previously reported risk genes for other neuropsychiatric conditions in both DNA sequencing and genome-wide association studies. We also show that these genes are enriched for several canonical biological pathways, suggesting early neurodevelopmental underpinnings of ADHD. Overall, this work provides critical new insight into the biology of ADHD and demonstrates the discovery potential of DNA sequencing in larger parent-child trio cohorts.

**Significance statement:** Given the important role of genetic factors in the development of attention-deficit/hyperactivity disorder (ADHD), research aimed at identifying risk genes can provide critical insight into underlying biological processes. We conducted whole-exome DNA sequencing in parent-child trios with ADHD, showing that these children have a significantly greater rate of rare and ultra-rare *de novo* gene-damaging mutations compared to unaffected controls, expanding our understanding of the genetic landscape of ADHD. We then use this information to identify *KDM5B* as a high-confidence risk gene for ADHD and highlight several enriched biological pathways. This work advances our etiologic understanding of ADHD and illustrates a previously unexplored path for risk gene discovery in this common neurodevelopmental disorder.

## Introduction

Attention-deficit/hyperactivity disorder (ADHD) is a common neurodevelopmental disorder in childhood (1) that places a significant burden on individuals, their families, and the community (2). ADHD is highly heritable (∼70-80%) (3), so identifying genes associated with the disorder will increase our understanding of underlying biological processes. Recent case-control genome-wide studies have identified ADHD risk loci by assessing common single-nucleotide polymorphisms (SNPs) through genome-wide association studies (GWAS) (4, 5). However, to date, SNP-heritability has only accounted for a small portion (∼15-30%) of the overall heritability estimates from twin studies, suggesting that other genetic factors, including rare genetic variants, may play an important role in ADHD risk (6). Indeed, previous studies have demonstrated that rare copy number variants (7) and very rare protein-truncating variants in evolutionarily constrained genes (8) are enriched in ADHD. Therefore, assessing rare variation may help identify potential ADHD risk genes. Despite previous research considering these different categories of genetic variation in ADHD, few specific high-confidence risk genes have yet been identified.

Studies of rare *de novo* genetic variants using parent-child trios have proven to be powerful for risk gene discovery in other neurodevelopmental disorders such as autism spectrum disorder (ASD) (9), developmental delay/intellectual disability (10), and Tourette’s disorder (11), leading to the discovery of risk genes. Since the background rate of *de novo* variants in the population is low, finding an elevated rate of damaging *de novo* variants suggests that we can leverage these variants to identify risk genes and underlying biological pathways. However, this approach has yet to be extensively leveraged for ADHD.

Only a few studies have used parent-child trios to investigate the genetics of ADHD. Studies examining *de novo* copy number variants (CNVs) suggest a greater rate of these variants in ADHD cases compared to rates in controls (12, 13), with the largest study suggesting a rate similar to that previously reported in ASD and Tourette’s disorder (13). However, given the large number of genes disrupted by CNVs, it is challenging to identify specific risk genes from these variants. Whole-exome DNA sequencing studies enable the identification of *de novo* sequence variants affecting single genes. There have been a few small whole-exome DNA sequencing studies of parent-child trios focused on ADHD (14-16), each identifying rare damaging *de novo* sequence variants in 11-30 parent-child trios, supporting the discovery potential of applying this approach in larger ADHD cohorts.

Here, we conducted whole-exome DNA sequencing in 152 parent-child trios (456 individuals in total), comprising a child with ADHD and both biological parents, to identify rare and ultra-rare *de novo* genetic variants and compare rates with previously sequenced controls without ADHD. For the first time, we demonstrate that rare and ultra-rare *de novo* protein-truncating variants (PTVs, including single nucleotide variants introducing premature stop codons, frameshift indels, and canonical splice site variants), as well as missense variants predicted to be damaging (Mis-D), are enriched in ADHD cases compared to controls.

Combining our results with a large independent case-control DNA sequencing study (3,206 ADHD cases and 5,002 typically developing controls) (8), we identify *lysine demethylase 5B* (*KDM5B)* as a high-confidence risk gene for ADHD (FDR<0.1). Finally, we identify overlap among genes harboring *de novo* damaging variants in ADHD and previously reported risk genes for other psychiatric conditions, and we conduct exploratory analyses to identify biological pathway enrichment. These new findings provide a critical step forward toward improving our etiologic understanding of ADHD, which may, in the future, inform the treatment of this common and impairing condition.

## Results

### Rare and ultra-rare de novo damaging variants are enriched in ADHD probands

We performed whole-exome DNA sequencing in 152 parent-child trios with ADHD collected from four sites (**Dataset S1**). We pooled this sequencing data and performed joint variant calling with whole-exome sequencing from 788 parent-child trios without ADHD, already sequenced as part of the Simons Simplex Collection. After applying our quality control methods, we compared rates of *de novo* variants in 147 ADHD parent-child trios and 780 control parent-child trios. Based on studies of other childhood-onset neuropsychiatric conditions (9, 11, 17, 18), we expected to find a greater rate of rare *de novo* damaging variants in ADHD probands versus controls. Damaging variants include protein-truncating variants (PTVs, including premature stop codons, frameshift, and splice site variants) and missense variants predicted to be damaging (Mis-D) by a “missense badness, PolyPhen-2, constraint” (MPC) score > 2 (19).

Results from this burden analysis demonstrate a greater rate of both rare and ultra-rare *de novo* damaging variants (PTVs + Mis-D) in ADHD cases versus unaffected controls (**Figure 1, Table S1, Dataset S2**). For rare *de novo* damaging variants (non-neuro gnomAD allele frequency < 0.001), the rate ratio was 1.55 (95% CI 1.02-2.30). We found a greater difference between cases and controls when narrowing our analysis to ultra-rare *de novo* damaging variants (non-neuro gnomAD allele frequency < 0.00005), with a rate ratio of 1.86 (95% CI 1.21-2.83, p=0.009) (**Table S1, Figure 1**). Within the subset of ultra-rare *de novo* damaging variants, we found a greater rate of PTVs (rate ratio 1.85, 95% CI 1.10-3.03, p=0.03) and a trend towards an increased rate of Mis-D variants in cases versus controls (rate ratio 1.91, 95% CI 0.78-4.36, p=0.12). As anticipated, we did not find differences in rates of *de novo* variants between cases and controls when including all (damaging and non-damaging) rare or all ultra-rare variants (**Table S1**).

**Figure 1:**
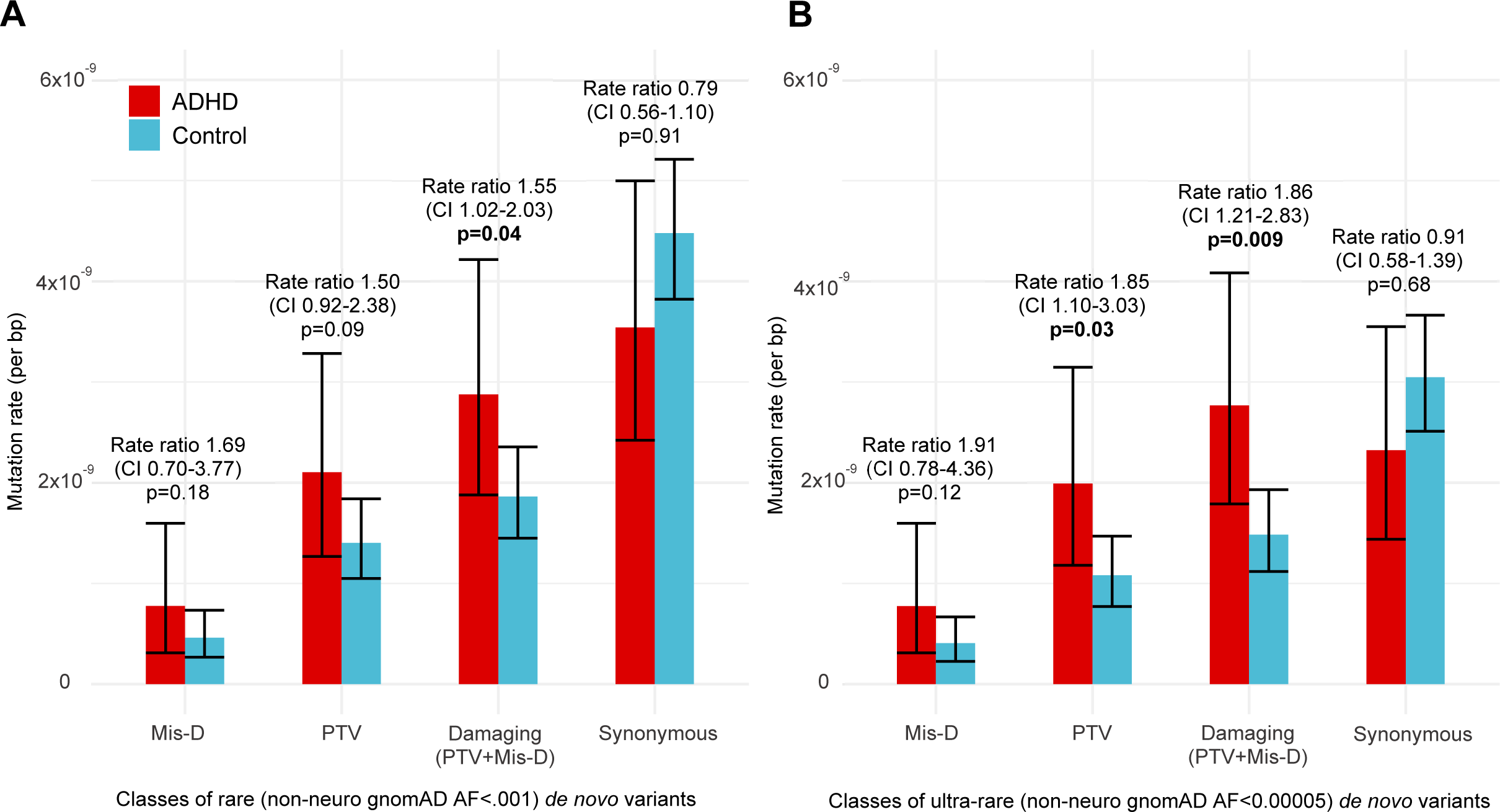
Rates of (A) rare and (B) ultra-rare *de novo* damaging mutations are enriched in ADHD probands (n=147) compared to controls (n=780). Rare variants have an allele frequency <0.001 (0.1%) in the non-neuro subset of the Genome Aggregation Database (gnomAD) and ultra-rare de novo variants have an allele frequency of <0.00005 (0.005%) in the non-neuro subset of gnomAD. The mutation rate per base pair (bp) includes only the “callable” loci in each family that meet sequencing depth and quality scores. Mutation rates are compared with a one-tailed rate ratio test with a p<0.05 considered significant. Bold p-values are significant. Error bars show 95% confidence intervals. PTVs, protein truncating variants, including frameshift, splice site, and stop-gain variants. Mis-D, missense variants predicted to be damaging with “missense badness, PolypPhen-2 constraint” (MPC) score>2.

### Recurrent ultra-rare damaging variants identify ADHD risk genes

Among 147 ADHD parent-child trios passing quality control, we identified 25 ultra-rare *de novo* damaging variants in 24 individuals (**Table 1, Dataset S2**). One gene, *KDM5B*, had two *de novo* PTVs in unrelated individuals in our ADHD trio cohort. To identify ADHD risk genes (genes harboring damaging variants more often than expected by chance), we combined our *de novo* parent-child trio findings with counts of ultra-rare PTVs and Mis-D (MPC > 2) variants identified in a large independent ADHD case-control dataset (3,206 ADHD cases and 5,002 typically developing controls) (8). Using this combined dataset, we applied the Transmission And De novo Association test (extTADA) (20) and identified *KDM5B* as a high-confidence risk gene (FDR=0.05), *POMT1* as a probable risk gene (FDR=0.21), and *YLPM1* as a probable risk gene (FDR=0.28) for ADHD (**Figure 2, Dataset S3**). This extTADA analysis estimates that 862 genes (95% CI 243-2,502) contribute to ADHD risk.

**Figure 2:**
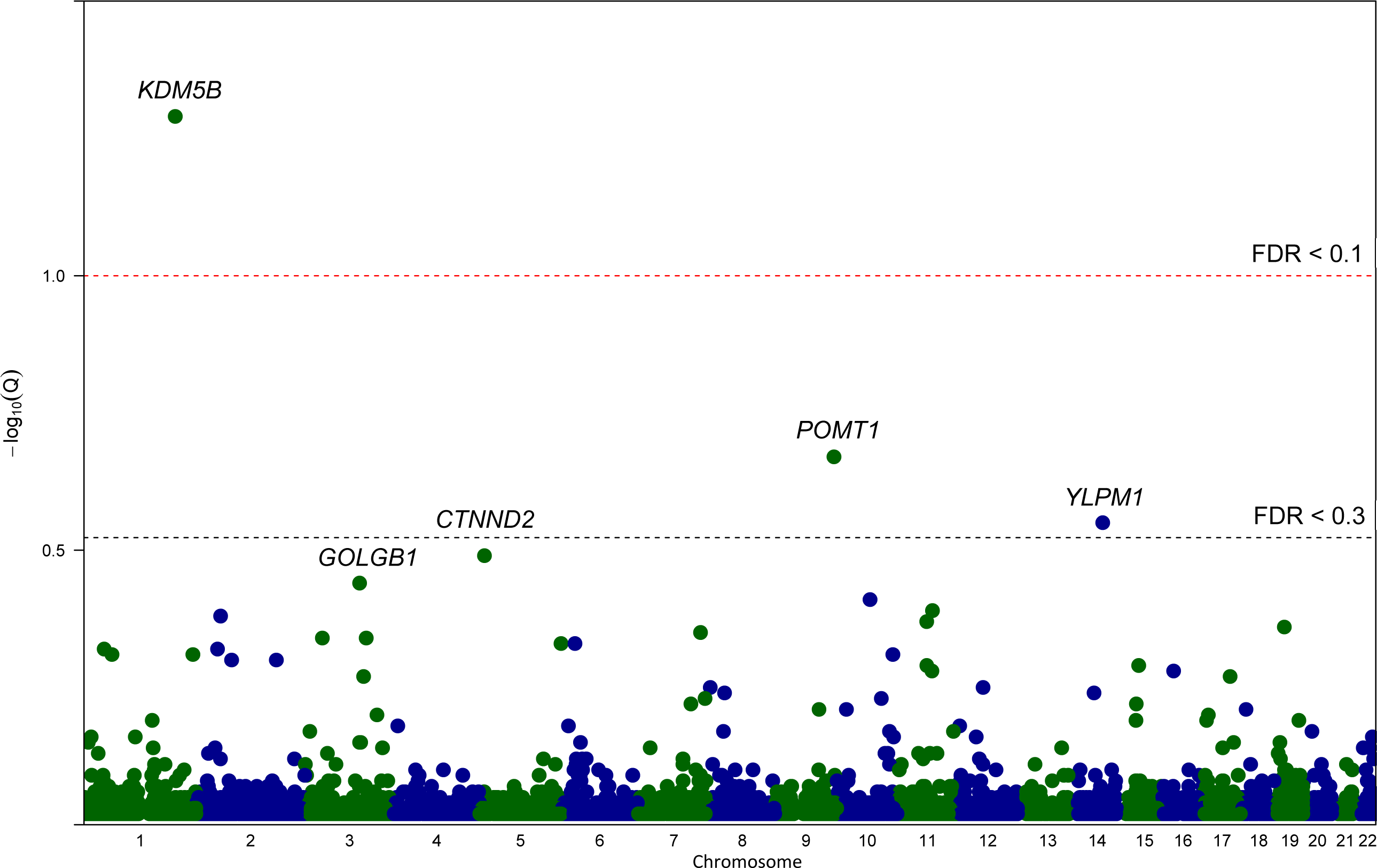
Gene-based test results for ADHD, combining the ultra-rare *de novo* damaging variants and independent case-control data. Results from extension of the Transmission And De novo Association test (extTADA) examining ultra-rare *de novo* protein-truncating variants and missense variants predicted to be damaging (MPC score>2) from the 147 ADHD parent-child trios and an independent group of 3,206 ADHD cases and 5,002 unaffected controls. Genes are organized by chromosome and the top 5 gene symbols are listed that have the lowest q values. Only one gene *KDM5B* is classified as a high-confidence risk gene (FDR, false discovery rate < 0.1) and two genes, *POMT1* and *YLPM1*, are classified as probable risk genes (FDR<0.3)

**Table 1:**
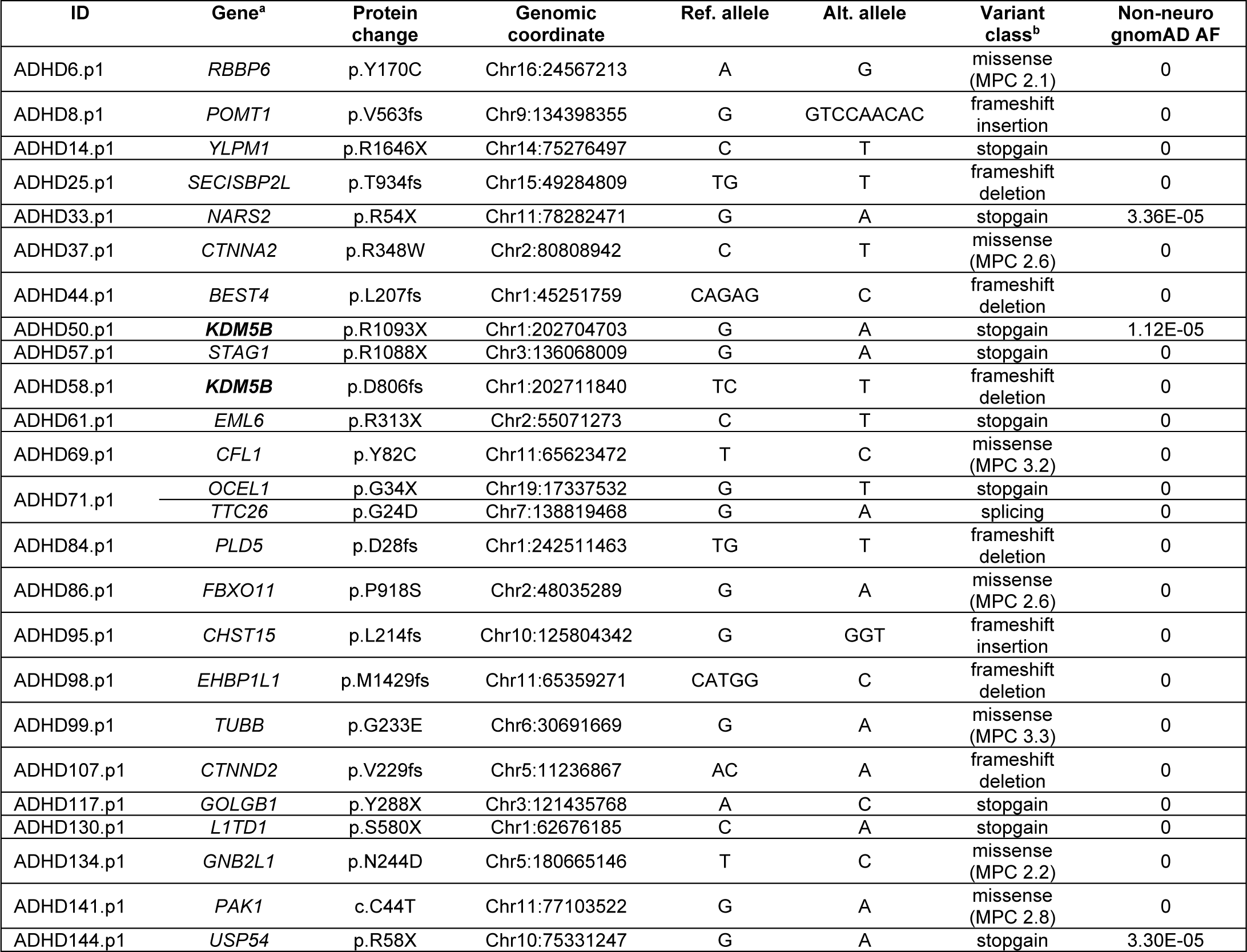
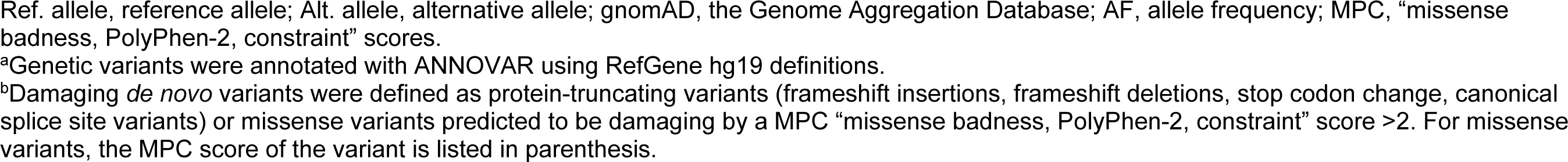
Ultra-rare *de novo* damaging variants identified in ADHD probands

### Genes with de novo damaging variants in ADHD overlap with risk genes for other psychiatric conditions

Using the list of 25 genes with ultra-rare *de novo* damaging variants (PTV and Mis-D) in 147 ADHD probands (**Table 1, Dataset S2**), we identified overlap with risk genes for other conditions (**Table S2**), using the Gene4Denovo database (21). *KDM5B* is also a risk gene for autism spectrum disorder (FDR=0), general developmental disorders (FDR=0), congenital heart disease (FDR=.005), complex motor stereotypies (FDR=0.06), and across all disorders in the Gene4Denovo database (FDR=0). *FBXO11* and *STAG1* were also both associated with developmental disorders (FDR= 0 and 0.00002, respectively) and across disorders (FDR=0 for both), and *PAK1* was a risk gene across disorders (FDR=.009). Additionally, we identified overlap between our list of 25 genes with ultra-rare *de novo* damaging variants in ADHD probands and gene-mapped loci from common variant GWAS studies in neuropsychiatric disorders in the GWAS Catalog (**Table S3**).

### Exploratory gene ontology and pathway enrichment

Using this same list of 25 genes harboring ultra-rare *de novo* damaging variants in ADHD trios, we also conducted exploratory analyses to identify enriched gene ontology and biological pathways. Several gene ontology and pathway-based sets were enriched for these 25 genes identified in ADHD (**Table S4**). The top pathway-based sets were CXCR4-mediated signaling events (q=0.004), Sema3A PAK dependent Axon repulsion (q=0.004), and ectoderm differentiation (q=0.008).

## Discussion

In this largest parent-child trio whole-exome DNA sequencing study of ADHD to date, we found a significantly greater rate of rare and ultra-rare *de novo* damaging variants in children with ADHD compared to unaffected controls (**Figure 1**). Combining our trio sequencing data with results from a large independent case-control DNA sequencing dataset, we identified *KDM5B* as a high-confidence risk gene and *POMT1* and *YLMP1* as two probable risk genes for ADHD (**Figure 2**).

Our sequencing data identified a 1.55-fold enrichment of rare *de novo* damaging variants in ADHD cases compared to unaffected controls (**Figure 1, Table S1**). Narrowing to ultra-rare *de novo* damaging variants increases this enrichment to 1.86-fold and strengthens statistical significance. It is important to note that these estimated enrichments have wide confidence intervals, so caution is warranted in interpreting these results, and replication in larger ADHD parent-child trio cohorts is needed. Nevertheless, our observed enrichment of rare *de novo* PTV and Mis-D variants is of a similar magnitude to enrichments reported in other neurodevelopmental disorders, including ASD and Tourette’s disorder (9, 11). This enrichment of rare and ultra-rare *de novo* damaging variants in ADHD cases compared to controls is also consistent with findings from the largest case-control DNA sequencing study that observed an enrichment of rare protein-truncating variants in constrained genes in ADHD cases, and this rate was similar in ASD cases (8). Finding an enrichment of rare *de novo* damaging variants in ADHD adds information about the genomic architecture of ADHD and supports the value of DNA sequencing studies in larger ADHD parent-child trio cohorts to identify risk genes in a manner which has led to the identification of over 100 high-confidence risk genes in ASD. The discovery potential of this approach is further reinforced by our estimate that 862 genes contribute to ADHD risk. Although the confidence interval of this estimate is wide and will need to be refined in larger cohorts, this finding further highlights a path toward systematic risk gene discovery in ADHD.

Our study identified ultra-rare *de novo* PTV variants in *KDM5B* in two unrelated individuals with ADHD (**Table 1, Dataset S2**). These individuals did not have diagnoses of ASD or intellectual disability. *KDM5B* is a histone-modifying enzyme that demethylates H3K4 and plays an important role in normal embryonic development (22), likely through epigenetic regulation of gene expression. This gene has been previously identified as a high-confidence risk gene for ASD (9) and developmental disorders more broadly (10). Interestingly, PTV mutations in *KDM5B* have also been reported in unaffected control subjects (8), and recessive mutations have been reported to cause a syndrome with developmental delay (23, 24). A review of the literature suggests that *KDM5B* likely causes an autosomal recessive developmental disorder, while dominant disease variants may exist (22). Our findings suggest, for the first time, that ADHD is included in the spectrum of phenotypic changes that may occur in the context of rare damaging variants in *KDM5B*.

We found several additional genes with *de novo* damaging variants in ADHD that are worth highlighting. First, we identified a *de novo* PTV in *POMT1*, which we identify as a probable risk gene for ADHD, based on FDR < 0.3 (**Table 1, Figure 2, Dataset S3**). POMT1 is a key enzyme in glycosylation of alpha-dystroglycan, and biallelic mutations are associated with dystroglycanopathies characterized by proximal muscular dystrophy (25) and often accompanied by intellectual disability (26). Second, we also identify *YLPM1* as a probable risk gene for ADHD. YLPM1 is involved in RNA binding and has been predicted to be involved in telomere maintenance, but to our knowledge, psychiatric manifestations related to *YLPM1* mutations have not been described previously. Third, although no other gene beyond *KDM5B* was found to have more than one ultra-rare *de novo* gene-damaging mutation in unrelated individuals using our definition of PTV and Mis-D, two additional genes, *CTNND2* and *EML6*, had a *de novo* PTV variant in one individual with ADHD and a *de novo* missense variant that was not predicted to be damaging (MPC<2) in an unrelated individual with ADHD (**Dataset S2**). Although not predicted to be damaging by MPC, the ultra-rare *de novo* missense variant in *CTNND2* was predicted to be damaging by PolyPhen2 (**Dataset S2**), and another PTV in *CTNND2* was identified in an ADHD case in the case-control dataset, but not in controls (**Dataset S3**). *CTNND2* encodes an adhesive junction protein, and mutations have been previously associated with intellectual disability in Cri-du-Chat syndrome, ASD, and epilepsy (27-29). Research suggests that *CTNND2* is important for the formation of dendritic spines and synapses (28). Finally, we identified individuals with ADHD who had *de novo* damaging variants in the genes *FBXO11* and *STAG1* (**Table 1**). These two genes have been previously identified as high-confidence risk genes for neurodevelopmental disorders in general (10). We did not see damaging variants in these genes in controls (**Datasets S2 and S3**). *FBXO11* encodes an F-box protein, and *de novo* variants have been associated with syndromic intellectual disability and behavioral difficulties, including ADHD (30, 31). *STAG1* encodes a component of cohesion involved with the separation of sister chromatids and has been associated with syndromic intellectual disability (32). In our study, ultra-rare damaging *de novo* variants in these genes were identified in children with ADHD who did not have intellectual disability or other known genetic syndromes. This highlights the potential range of clinical manifestations that may occur due to *de novo* damaging variants in these genes and suggests potential clinical implications for identifying *de novo* damaging variants.

Genes harboring rare *de novo* gene-damaging variants in the ADHD cases not only overlapped with high-confidence risk genes identified in previous DNA sequencing studies of other neuropsychiatric conditions (**Table S2**) but also overlapped with genes mapped from genome-wide significant common variants identified in previous GWA studies (**Table S3**). Although there was no overlap with the 76 prioritized risk genes identified in the recent large ADHD GWAS (4), there was overlap between genes mapped from externalizing-related disorders more broadly (33). These findings add to the growing evidence supporting the convergence of common and rare variants in ADHD (4) and psychiatric disorders in general (9, 34).

Finally, we conducted exploratory ontology and pathway analyses of genes harboring *de novo* damaging variants in our ADHD cases. In interpreting these results, it is important to note that many of these genes may not be true ADHD risk genes and replication of these exploratory findings are needed as more high-confidence risk genes are identified. Nevertheless, we observed a significant enrichment of several biological processes. Of note, one of the top pathways is ectoderm differentiation (**Table S4**), suggesting early neurodevelopmental underpinnings of ADHD. In the largest recent GWAS study of ADHD, gene-linked loci were enriched for expression in early brain development (4), also suggesting the possible role of early embryonic changes in the development of ADHD.

This study has limitations that should be considered. For comparing mutation rates, the ideal controls would have been sequenced simultaneously with cases and assessed for ADHD. This study prioritized sequencing ADHD parent-child trios and used controls that had been previously sequenced using similar methods and scored in the normal range of the ADHD subscale of the Child Behavioral Checklist (CBCL) or the Adult Behavioral Checklist (ABCL). We attempted to minimize batch effects by focusing on the intersection of the capture platforms as done in other DNA sequencing studies (11, 18, 35). Another limitation is that our study focused on the coding region of the genome, and it is possible that important rare variants also occur in the noncoding region. Currently, understanding the biological and clinical relevance of non-coding variation remains challenging, but future studies of ADHD may utilize whole-genome sequencing technologies.

Despite these limitations, our results are important by demonstrating, for the first time, an enrichment of rare and ultra-rare *de novo* damaging variants in ADHD cases compared to unaffected controls and identifying *KDM5B* as a high-confidence risk gene for ADHD. These findings reinforce the value of DNA sequencing of parent-child trios in larger cohorts to identify additional high-confidence risk genes for ADHD. Identifying risk genes that can be studied in model systems may offer additional insight into the underlying biology of ADHD and has the potential to inform clinical care for individuals and families.

## Materials and Methods

### Participants

This study was approved by the local institutional review boards of all participating institutions and informed consent/assent was obtained from all participants. A total of 152 parent-child trios (456 individuals in total), comprising a child meeting DSM-IV or DSM-5 criteria for the diagnosis of ADHD and both biological parents, were included in this study. Trios were identified from four sites: the University of São Paulo School of Medicine (n=30), the Centre for Addiction and Mental Health in Toronto (n=37), Florida International University (n=13), and the Genizon biobank from Génome Québec (n=72). All subjects were assessed by structured clinical interviews. Exclusion criteria included a diagnosis of ASD, intellectual disability, psychosis, mood disorders (including bipolar disorder), and clinically significant medical or neurological disease. We prioritized the study of simplex (no known family history of ADHD) parent-child trios to increase the likelihood of detecting *de novo* variants. Control subjects were 788 unaffected parent-child trios, selected from the Simons Simplex Collection from the National Institutes of Health Data Archive (https://nda.nih.gov/edit_collection.html?id=2042) (36). Control subjects did not have ASD and were selected to be in the normal range for the attentional problems subscale from the CBCL or the ABCL (t score < 64.5), which has been shown to predict ADHD diagnosis (37).

### Whole-exome DNA sequencing

Exome capture and whole-exome DNA sequencing of DNA from 80 children with ADHD and their parents were conducted at the Yale Center for Genomic Analysis (YCGA) using the IDT xGen V1 capture and the Illumina NovaSeq6000 sequencing instrument. An additional 72 ADHD parent-child trios were sequenced by Genome Quebec using the Agilent SureSelect All Exon V7 capture and the Illumina NovaSeq6000 sequencing instrument. 788 control parent-child trios were previously sequenced as part of the Simons Simplex Collection, using the NimbleGen SeqCap EzExomeV2 capture and the Illumina HiSeq 2000 sequencing instrument. We performed joint variant calling with sequencing data from all cases and controls (940 trios, 2,820 individuals in total).

### Sequencing alignment and variant identification

Alignment and variant calling of the DNA sequencing reads followed the Genome Analysis Toolkit (GATK) best practice guidelines (38) as previously described by our group (35). To minimize potential downstream effects of differential coverage between the different capture platforms, a target bed file was created using the intersection of target regions of the three capture platforms (IDT xGen V1, Agilent SureSelect All Exon V7, and SeqCap EzExome V2). Case and control samples were called jointly using GATK GenotypeGVCF tools and variant score recalibration was applied to all called variants. Passing variants were then annotated using the RefSeq hg19 gene definitions and external databases using ANNOVAR (39).

### Quality control of de novo variants

Parent-child trios were excluded if unexpected family relationships were identified using relatedness statistics (40). Trios were also omitted if the children were observed to have an outlier number of *de novo* variants (>20). PLINK/SEQ istats was used to generate quality control statistics for both cases and controls, and principal component analyses were used to remove outliers from the analysis (see **Figure S1** and **Dataset S1** for details). After these quality control steps, we analyzed 147 parent-child trios with ADHD and 780 control parent-child trios for *de novo* variants.

As previously described (35), we then used stringent thresholds to assess *de novo* mutations. This included that the child was heterozygous for the variant with an alternate allele frequency between 0.3 and 0.7 in the child and < 0.05 in the parents, sequencing depth of ≥ 20 in all family members at the variant position, alternate allele depth ≥ 5, mapping quality ≥ 30. Calls were limited to one variant per person per gene, retaining variants with the most severe consequence (9). We filtered to include rare *de novo* variants with an allele frequency < 0.001 (0.1%) in the “non-neuro” subset of the Genome Aggregation Database (gnomAD v2.2.1). Within this rare set of *de novo* variants, we defined an ultra-rare subset, defined as having an allele frequency of < 0.00005 in the non-neuro subset of gnomAD (41). The gnomAD v2.2.1 non-neuro dataset contains exome sequencing data from 104,068 individuals who were not ascertained for having a neurologic or psychiatric condition in case-control studies.

### Mutation rate analysis

We calculated the rate of *de novo* variants per base pair in cases and controls. The GATK DepthofCoverage tool was used to determine the denominator of the “callable” base pairs per family. We required callable bases to have a sequencing depth of ≥ 20x in all family members and mapping quality ≥ 30. We limited our analyses to variants in the callable exome to further minimize potential calling bias between cases and controls. Mutation rates were divided by 2 to calculate haploid rates, and confidence intervals were calculated (pois.conf.int, pois.exact function from epitools v0.5.10.1 in R). We used a one-tailed rate ratio test to compare *de novo* mutation rates between cases and controls (rateratio.test v1.1 in R). Based on studies of other childhood-onset neuropsychiatric conditions (9, 11, 17, 18), we hypothesized that rare and ultra-rare *de novo* PTV variants and missense variants predicted to be damaging (Mis-D) would be enriched in cases compared to controls. Mis-D variants were identified using the integrated “missense badness, PolyPhen-2, constraint” (MPC) score > 2 (19) as done in other recent studies (8, 18, 42). The combined group of *de novo* PTV and Mis-D variants were considered “damaging” variants.

### Transmission and De Novo Association Test Analysis

We used a Bayesian extension of the original Transmission And De novo Association test (extTADA) (20) to integrate *de novo* and case-control variants in a hierarchical model to increase the power of identification of risk genes for ADHD. We obtained mutation counts for PTVs and Mis-D variants (MPC > 2) from an independent case-control study including 3,206 individuals with ADHD and 5,002 typically developing controls (8). These individuals did not have diagnoses of autism, intellectual disability, schizophrenia, bipolar disorder, affective disorders, or anorexia (8). We ran extTADA to calculate the Bayes factor and q-values (false discovery rate, FDR) for each gene (details in Supplemental methods) (**Dataset S3**). We applied commonly used statistical thresholds to define “probable” (FDR < 0.3) and “high confidence” (FDR < 0.1) risk genes (17).

### Gene set overlap

We examined if our list of genes with ultra-rare *de novo* damaging variants (PTV or Mis-D) in the ADHD probands overlapped with genes implicated in other DNA sequencing studies and genome-wide association studies. The Gene4Denovo database (21) (http://www.genemed.tech/gene4denovo/home) integrates *de novo* mutations from 68,404 individuals across 37 different phenotypes, including several neuropsychiatric conditions, but not including ADHD. We assessed the overlap between the Gene4Denovo candidate gene list (release version updated 07/08/2022) with FDR < 0.1 and our list of genes with ultra-rare damaging *de novo* variants. The GWAS Catalog (43, 44) was used to examine if this same list of genes harboring *de novo* damaging variants overlapped with loci mapped to genes in previous genome-wide association studies of neuropsychiatric phenotypes. The GWAS Catalog identifies past studies through weekly PubMed searches and extracts data for SNPs with p < 1 x 10^-5^ in the overall (initial GWAS + replication) population. All curated trait descriptions in the GWAS Catalog are mapped to terms from the Experimental Factor Ontology (EFO), which provide a systematic description of traits to support the annotation, analysis, and visualization of data. We limited our overlap analysis to traits in the GWAS Catalog that were categorized under the umbrella terms ‘nervous system disease’ or ‘psychiatric disorder’ (additional details found at https://www.ebi.ac.uk/gwas//docs).

### Exploratory pathway analysis

We used ConsensusPathDB (45) (http://cpdb.molgen.mpg.de/, Latest Release 35, 05.06.2021) to assess if the list of genes harboring rare *de novo* damaging variants in ADHD probands were over-represented in gene-ontology based sets and biological pathways. This network interaction tool integrates information from 31 public resources. The following default settings were used for the gene set over-representation analysis with gene identifier type of Ensembl; Pathway based sets: pathways as defined by pathway databases, select all resources, minimum overlap with input list=2, p-value cutoff=0.01; Gene ontology categories: gene ontology levels 2, 3, 4, and 5 categories, select all categories, p-value cutoff=0.01.

## Data Availability

All data produced in the present work are contained in the manuscript.

## Acknowledgments

We thank all of the families who contributed to this research study. We would also like to thank Kyle Satterstrom, Anders Børglum, and Mark Daly for sharing their case-control PTV and Mis-D counts for this analysis and all of the team members who contributed to data collection, including Chelsea Dale and Juliana Acosta. This work was supported by a Klingenstein Third Generation Foundation ADHD Fellowship grant (E.O.), a Yale Child Study Center Faculty Development Award (T.V.F.), and the Allison Family Foundation (T.V.F.). Brazilian samples were recruited and collected with support from São Paulo Research Foundation (FAPESP, grant 2016/22455-8). E.O. was supported by the National Institute of Mental Health grants R25MH077823, T32MH018268, and K08MH128665. L.C.F. was supported by grant #2021/08540-0, São Paulo Research Foundation (FAPESP). J.P. was supported by the Bradley Hospital COBRE Center for Sleep and Circadian Rhythms in Child and Adolescent Mental Health (P20GM139743, PI Carskadon). Seventy-two of the sequenced ADHD parent-child trios were from the Génome Québec Genizon Biobank. The content is solely the responsibility of the authors and does not necessarily represent the official views of the National Institutes of Health. Simons Simplex Collection (SSC) control parent-child trio genetic data used in the preparation of this manuscript were obtained from the NIH-supported National Database for Autism Research (NDAR). NDAR is a collaborative informatics system created by the National Institutes of Health to provide a national resource to support and accelerate research in autism. Dataset identifier: https://nda.nih.gov/edit_collection.html?id=2042. This manuscript reflects the views of the authors and may not reflect the opinions or views of the NIH or of the Submitters submitting original data to NDAR. We are grateful to all of the families at the participating SSC sites, as well as the principal investigators (A. Beaudet, R. Bernier, J. Constantino, E. Cook, E. Fombonne, D. Geschwind, R. Goin-Kochel, E. Hanson, D. Grice, A. Klin, D. Ledbetter, C. Lord, C. Martin, D. Martin, R. Maxim, J. Miles, O. Ousley, K. Pelphrey, B. Peterson, J. Piggot, C. Saulnier, M. State, W. Stone, J. Sutcliffe, C. Walsh, Z. Warren, E. Wijsman). We appreciate obtaining access to phenotypic data on SFARI Base. Approved researchers can obtain the SSC population dataset described in this study (https://www.sfari.org/resource/simons-simplex-collection/) by applying at https://base.sfari.org.

## Supporting Information for

**Supporting Information Text**

**Details of Transmission and De Novo Association Test Analysis**:

We ran extTADA following the code outlined at https://github.com/hoangtn/extTADA/blob/master/examples/extTADA_OneStep.ipynb (1). Unlike the original TADA, extTADA uses a Markov chain Monte Carlo (MCMC) approach to calculate all parameters that are used as input in the traditional TADA (2) through sampling from the posterior in one step with resulting credible intervals. Parameter estimation led to the following estimates of (1) proportion of risk genes (π) (lower-upper credible intervals): 4.40% (1.24% – 12.79%); (2) average relative risk (γ) (lower-upper credible intervals): misD DN = 13.96 (1.22 – 67.51), PTV DN = 20.42 (3.27 – 66.51), misD CC = 1.60 (1.0 – 4.63), PTV CC = 1.74 (1.12 – 5.52); and (3) variability in relative risk estimates per gene (β) (lower-upper credible intervals): misD DN = 0.83, PTV DN = 0.82, misD CC = 5.65, PTV CC = 3.54. These parameters were used by the extTADA function to calculate the Bayes factor and q-values (FDR) for each gene.

For the calculation of absolute number of ADHD risk genes, we multiplied the total number of genes included in the extTADA analysis (19,560) by the proportion of risk genes estimated by the extTADA pipeline. All genes from the list generated by denovolyzeR(3) except for American College of Medical Genetics genes (*ACTA2, ACTC1, APC, APOB, ATP7B, BMPR1A, BRCA1, BRCA2, CACNA1S, COL3A1, DSC2, DSG2, DSP, FBN1, GLA, KCNH2, KCNQ1, LDLR, LMNA, MEN1, MLH1, MSH2, MSH6, MUTYH, MYBPC3, MYH11, MYH7, MYL2, MYL3, NF2, OTC, PCSK9, PKP2, PMS2, PRKAG2, PTEN, RB1, RET, RYR1, RYR2, SCN5A, SDHAF2, SDHB, SDHC, SDHD, SMAD3, SMAD4, STK11, TGFBR1, TGFBR2, TMEM43, TNNI3, TNNT2, TP53, TPM1, TSC1, TSC2, VHL*) were included in the exTADA analysis.

**Fig. S1.**
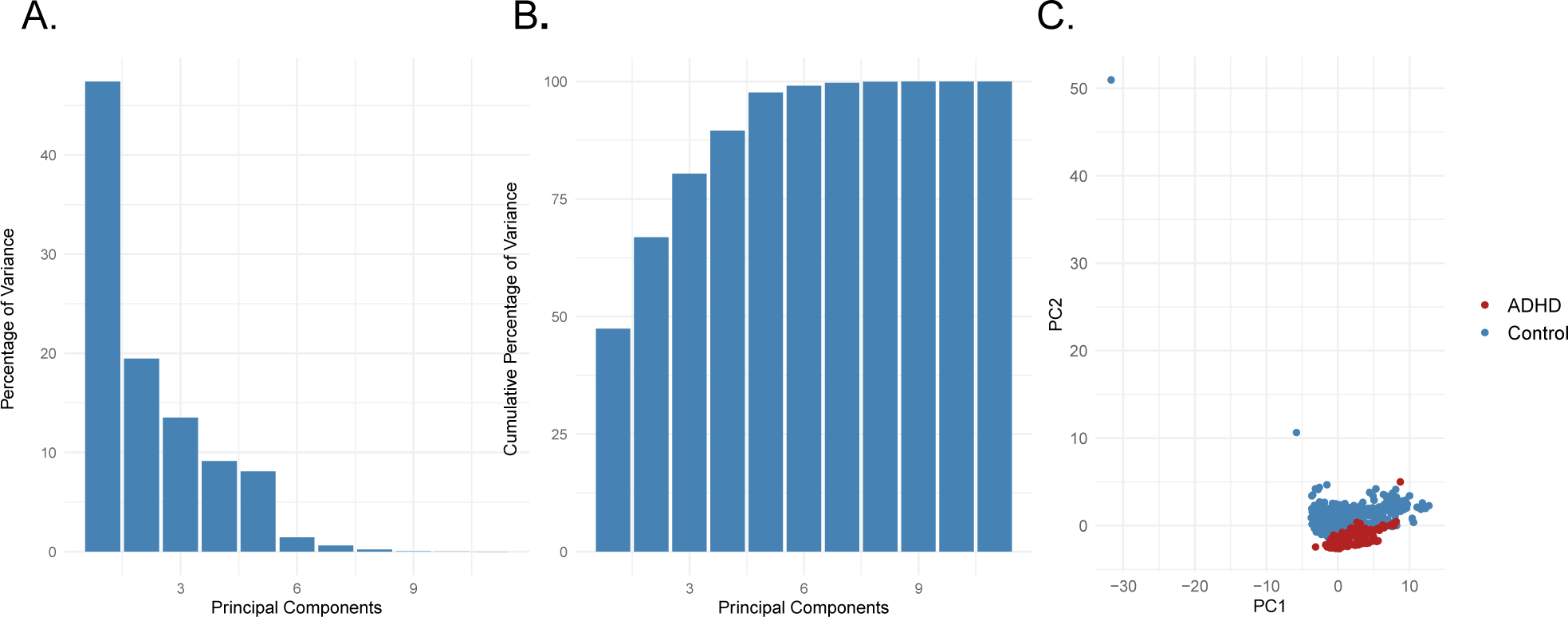
Plots from the principal components analysis (PCA). (A) Shows the percentage of variance captured by the 11 principal components from the exome metrics data from cases and controls. (B) Shows the cumulative percentage of variance captured by these components and demonstrates that over 75% of the cumulative variance was captured by the first 3 principal components. (C) Shows the first two principal components based on the PCA of the exome sequencing quality metrics. ADHD cases are plotted in red and controls in blue. This figure includes PCA outliers (>5 standard deviations in PCAs 1-3), which were removed during the quality control.

**Table S1.**
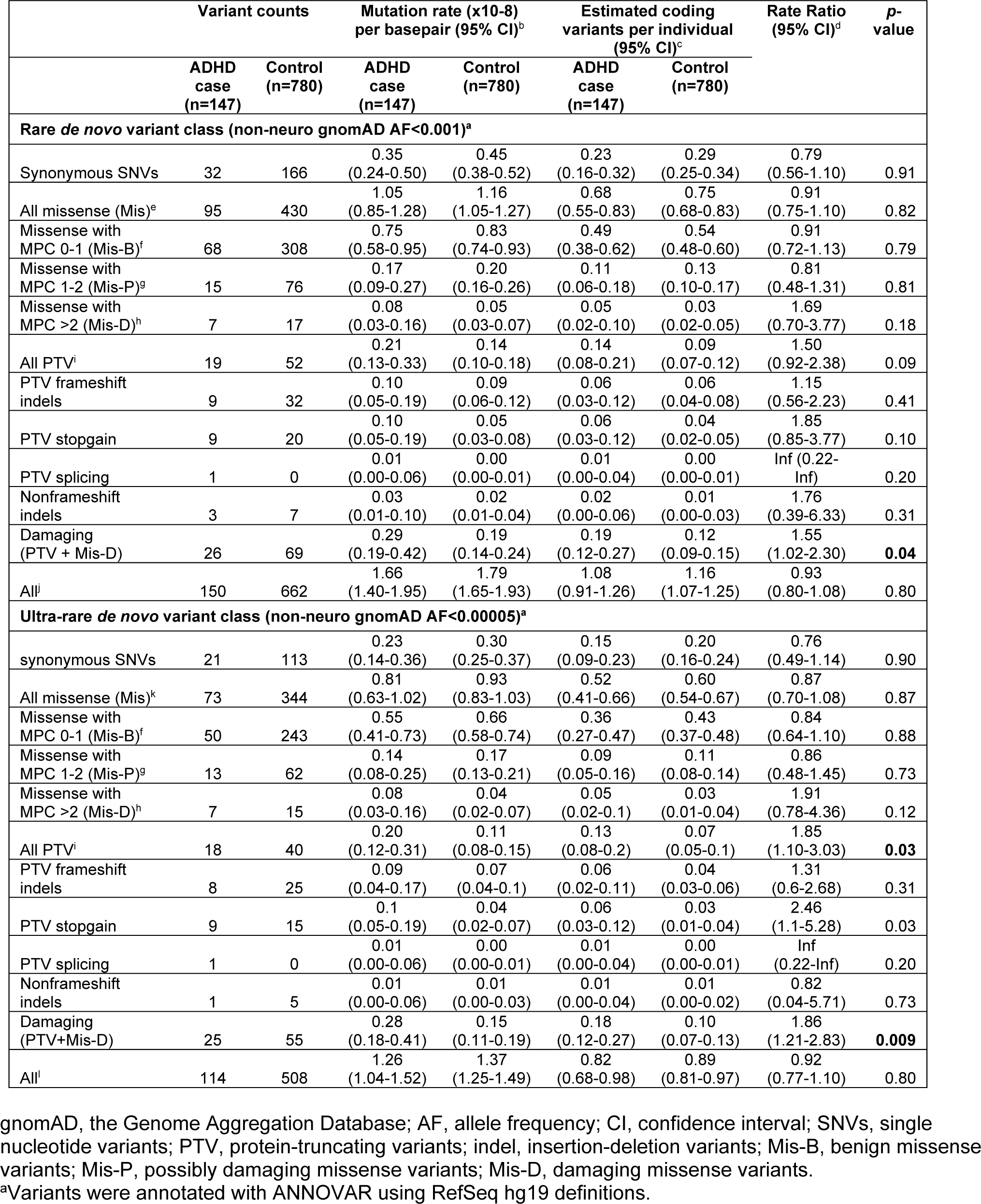
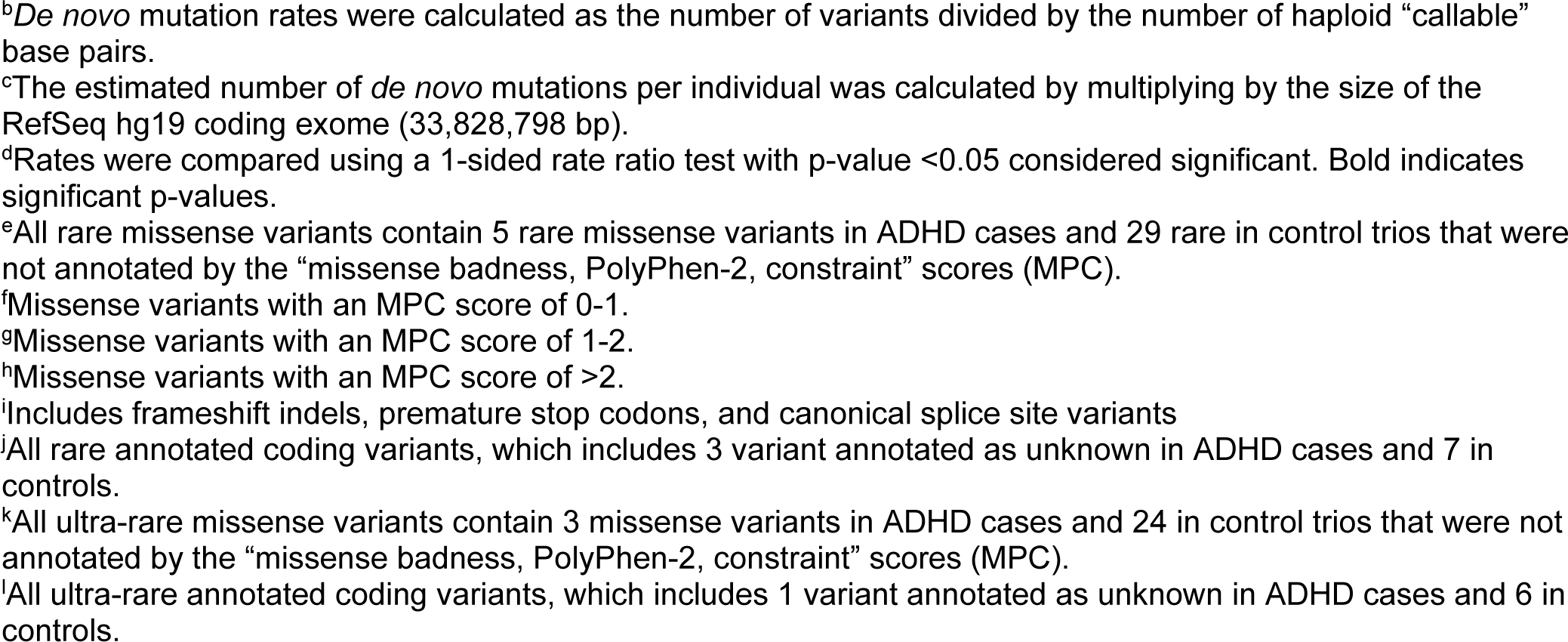
Distribution of classes of rare and ultra-rare *de novo* variants in ADHD cases and controls

**Table S2.**
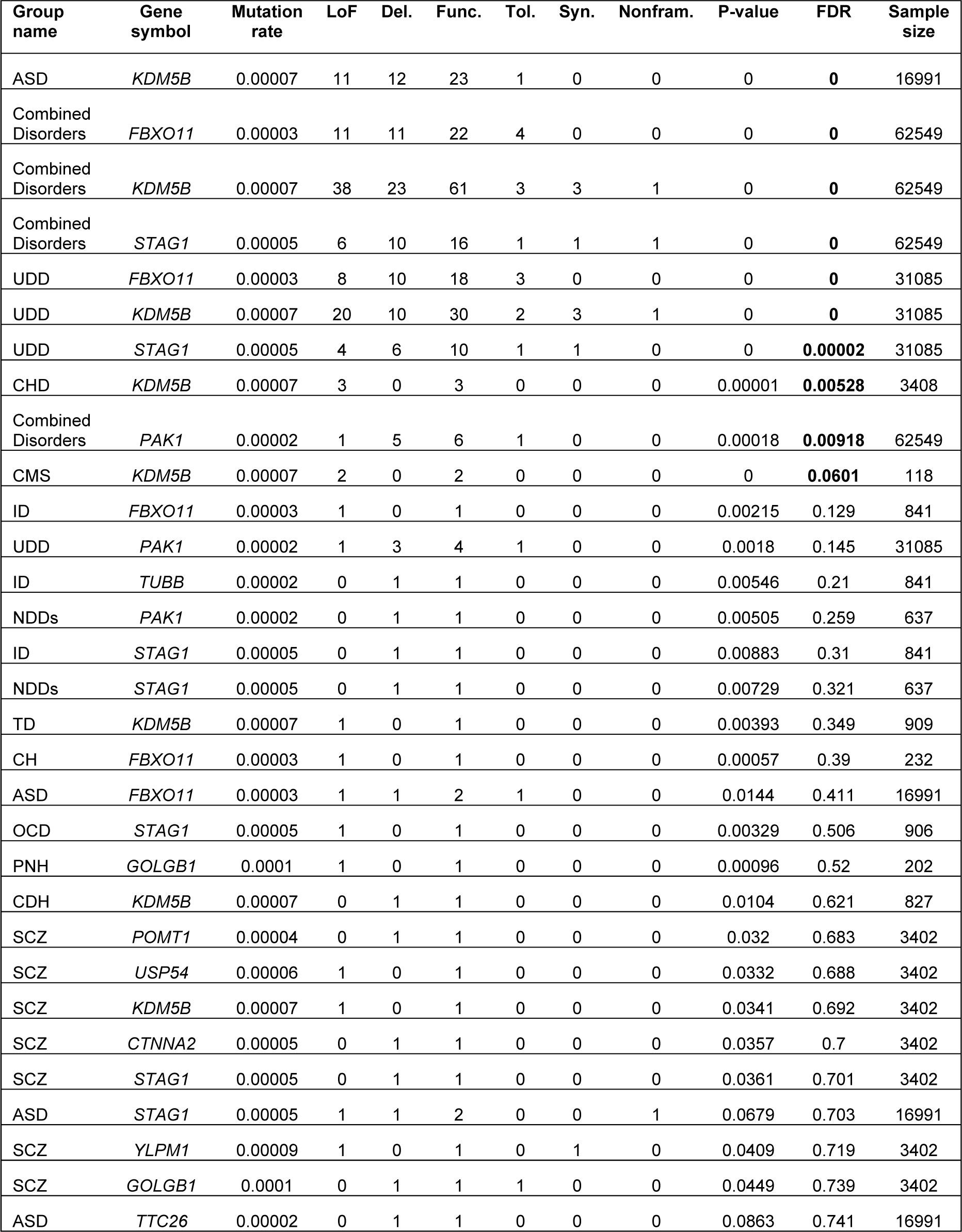
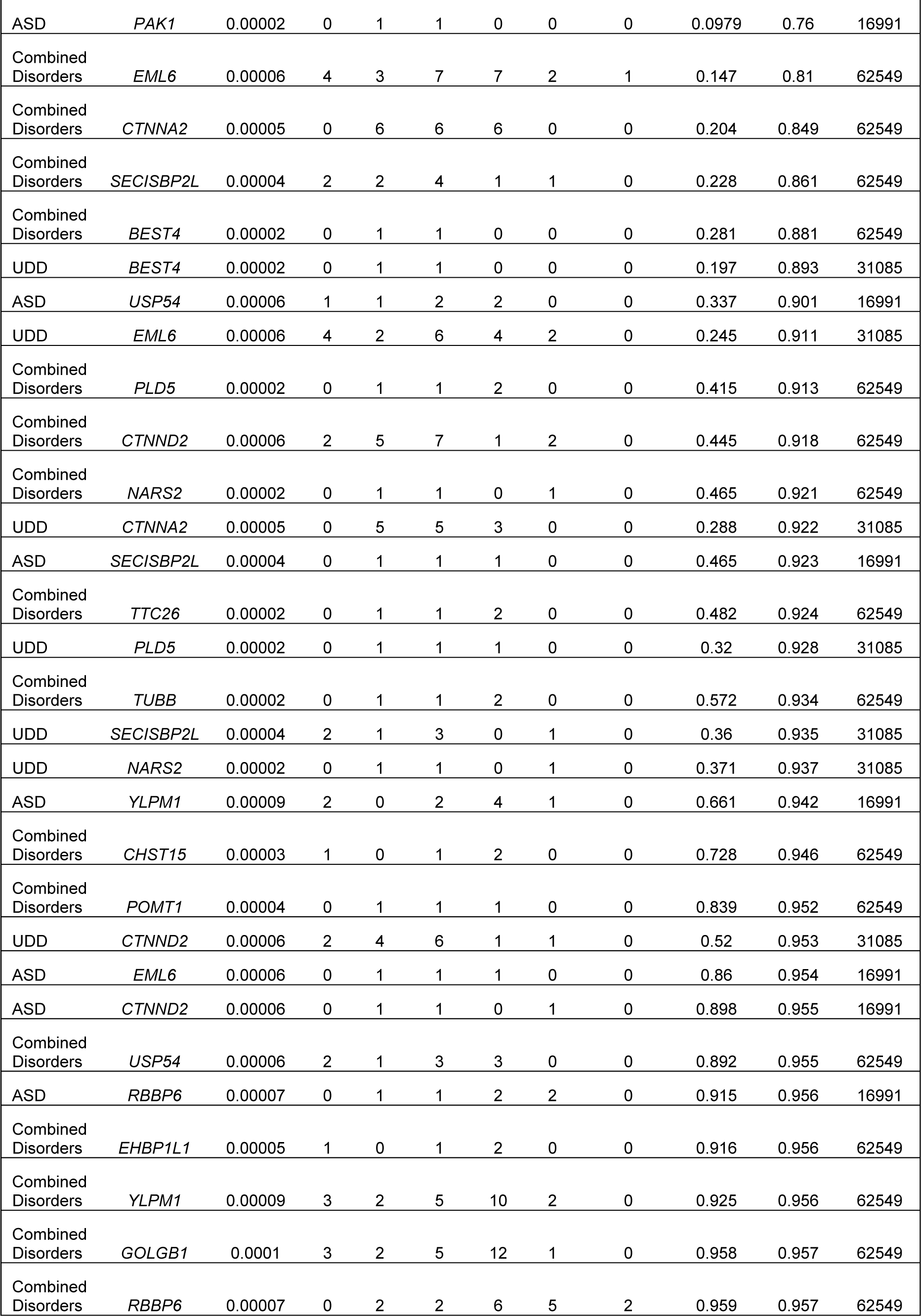
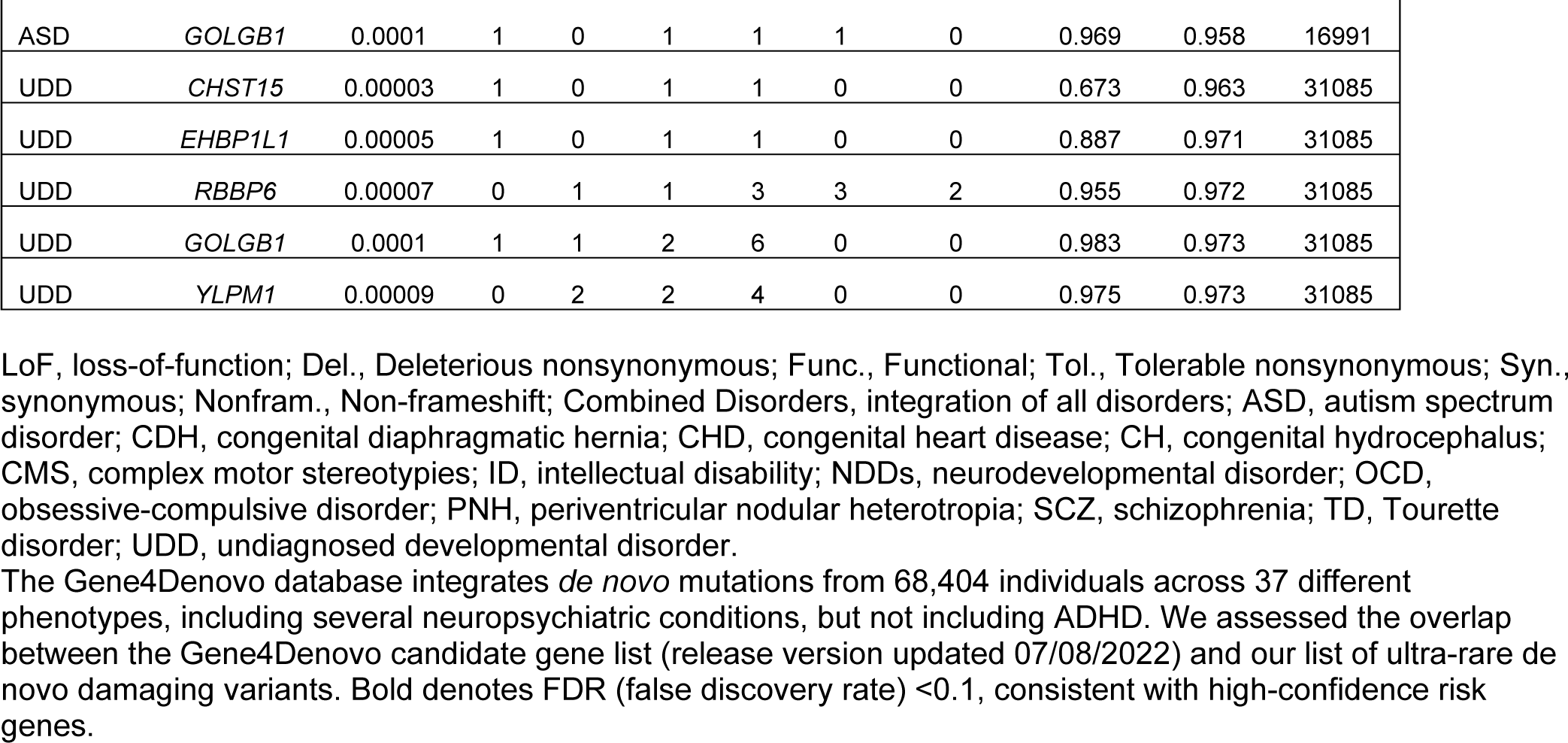
Overlap between genes harboring ultra-rare *de novo* damaging variants (PTV + Mis-D) in ADHD probands and genes identified in other DNA sequencing studies of parent-child trios using the Gene4Denovo database

**Table S3.**
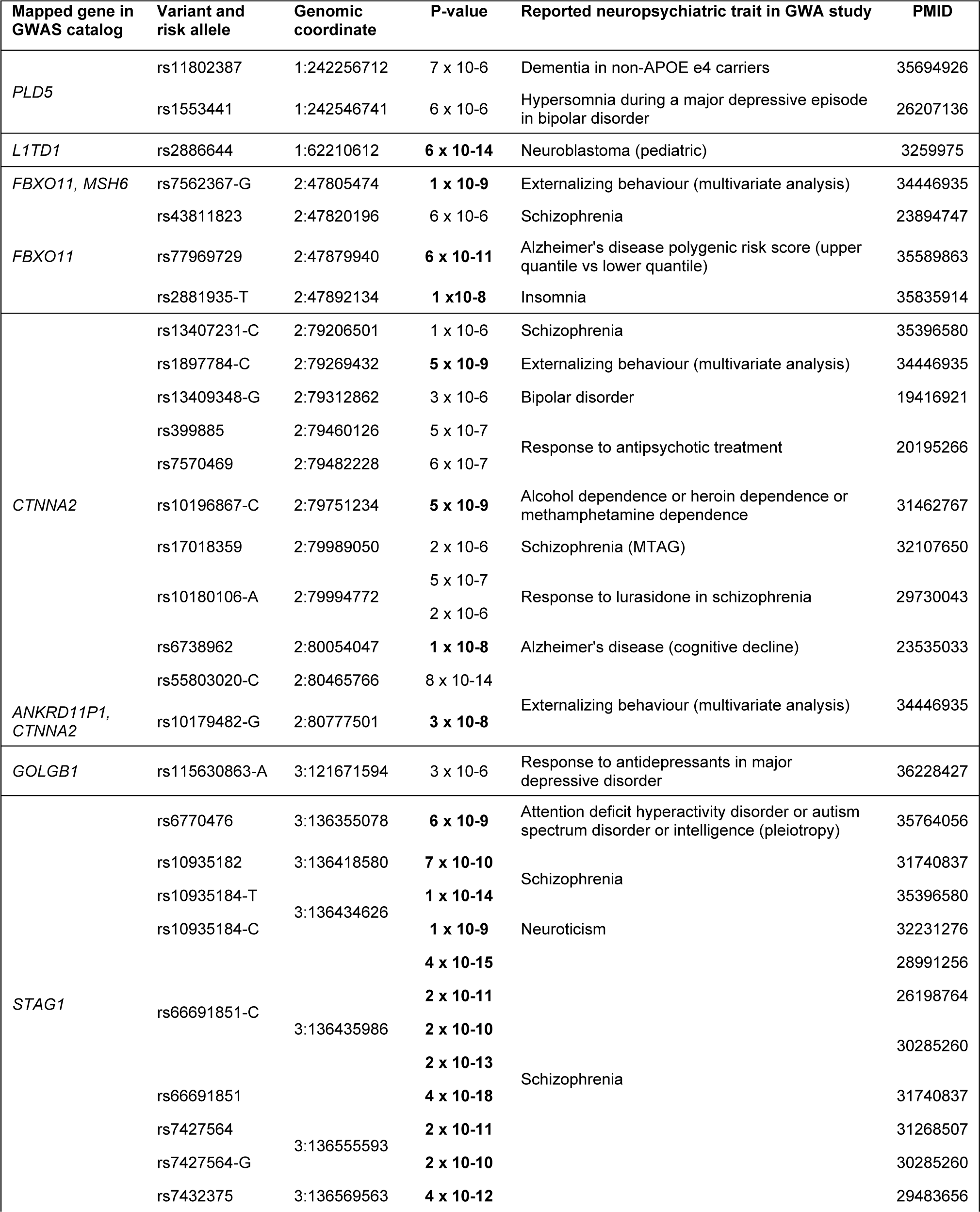
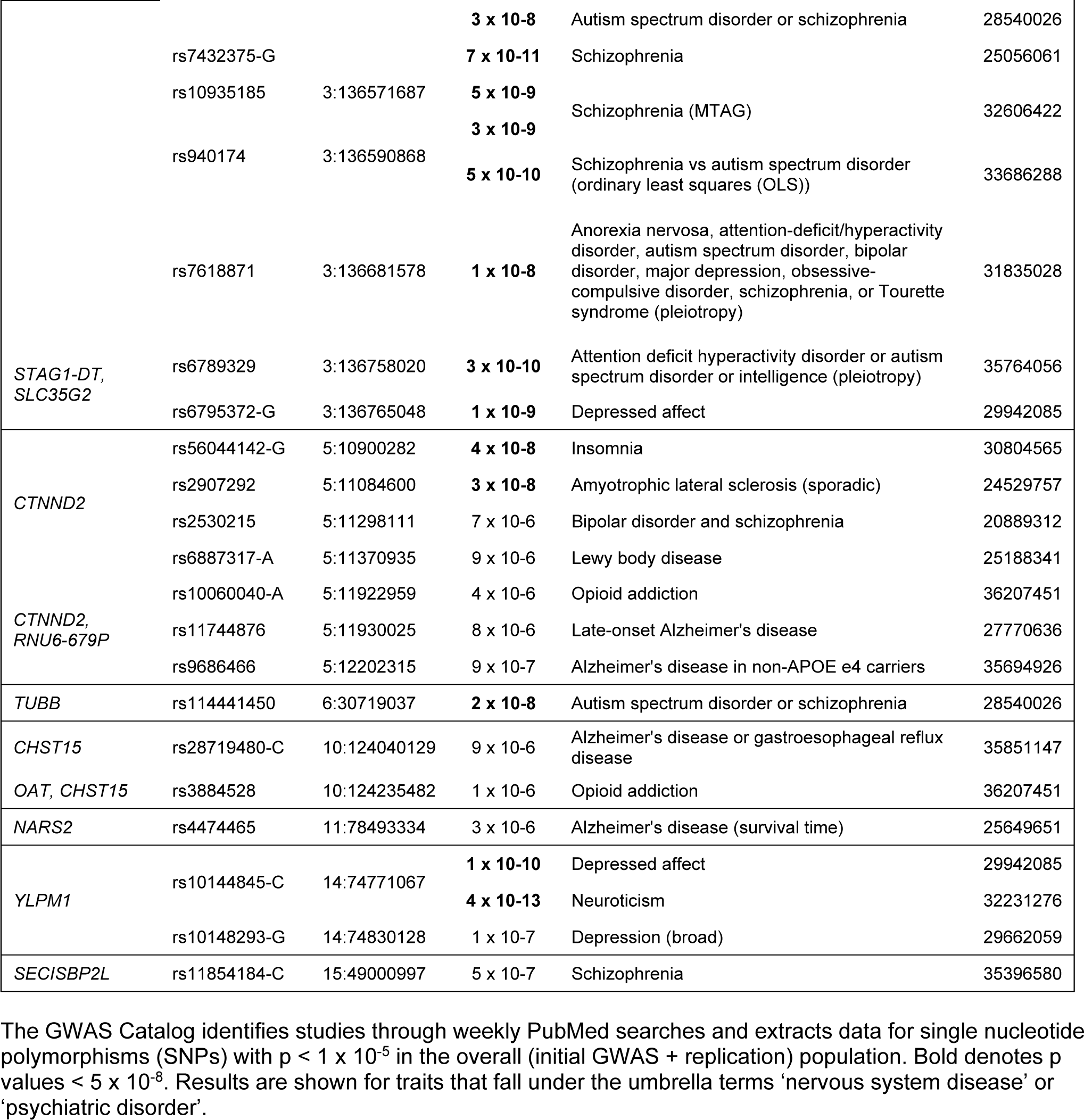
Overlap between the genes harboring ultra-rare de novo damaging variants (PTV or Mis-D) in ADHD probands and loci mapped to genes in genome-wide association studies (GWAS) of neuropsychiatric conditions using the GWAS Catalog

**Table S4:**
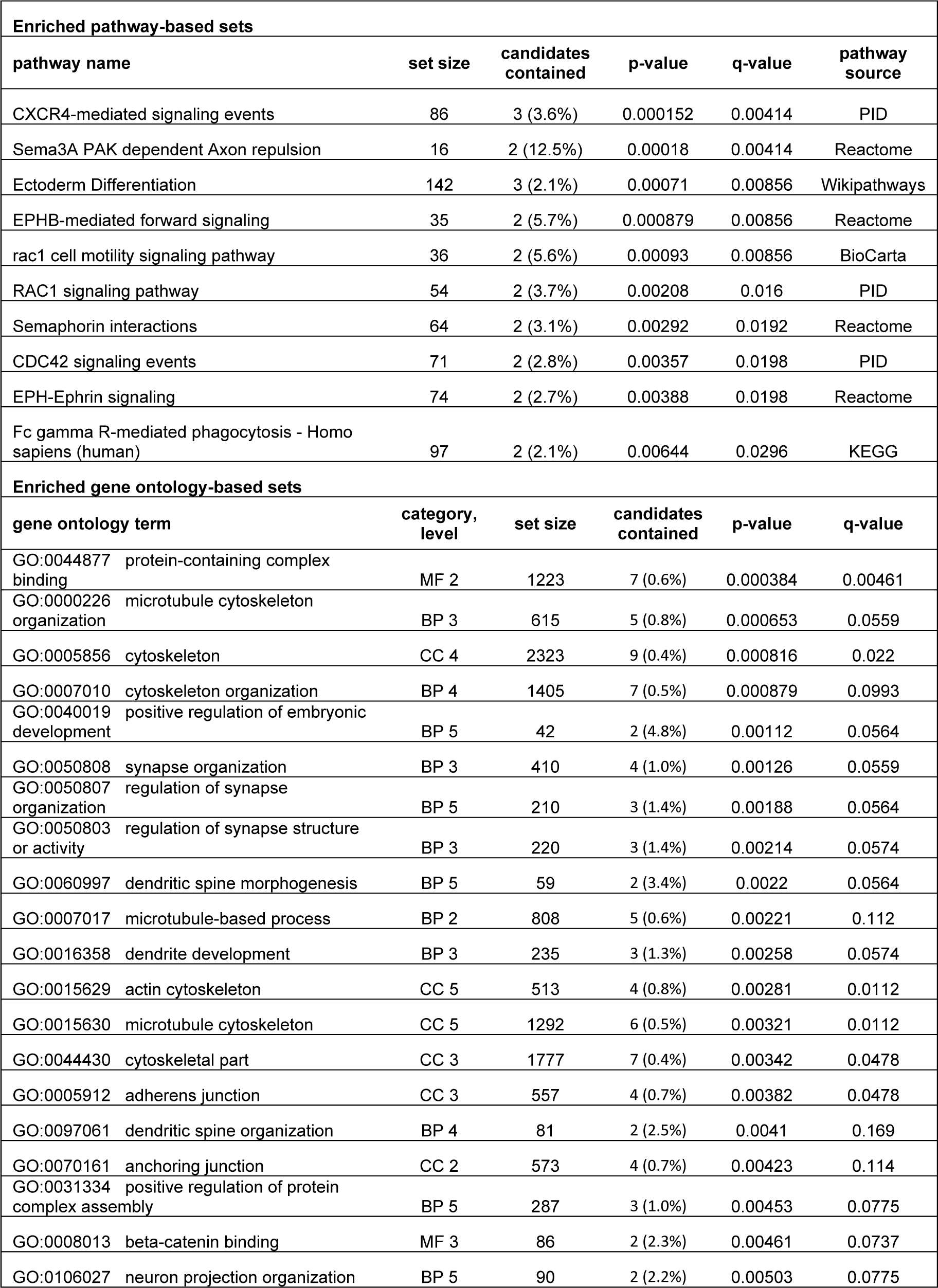
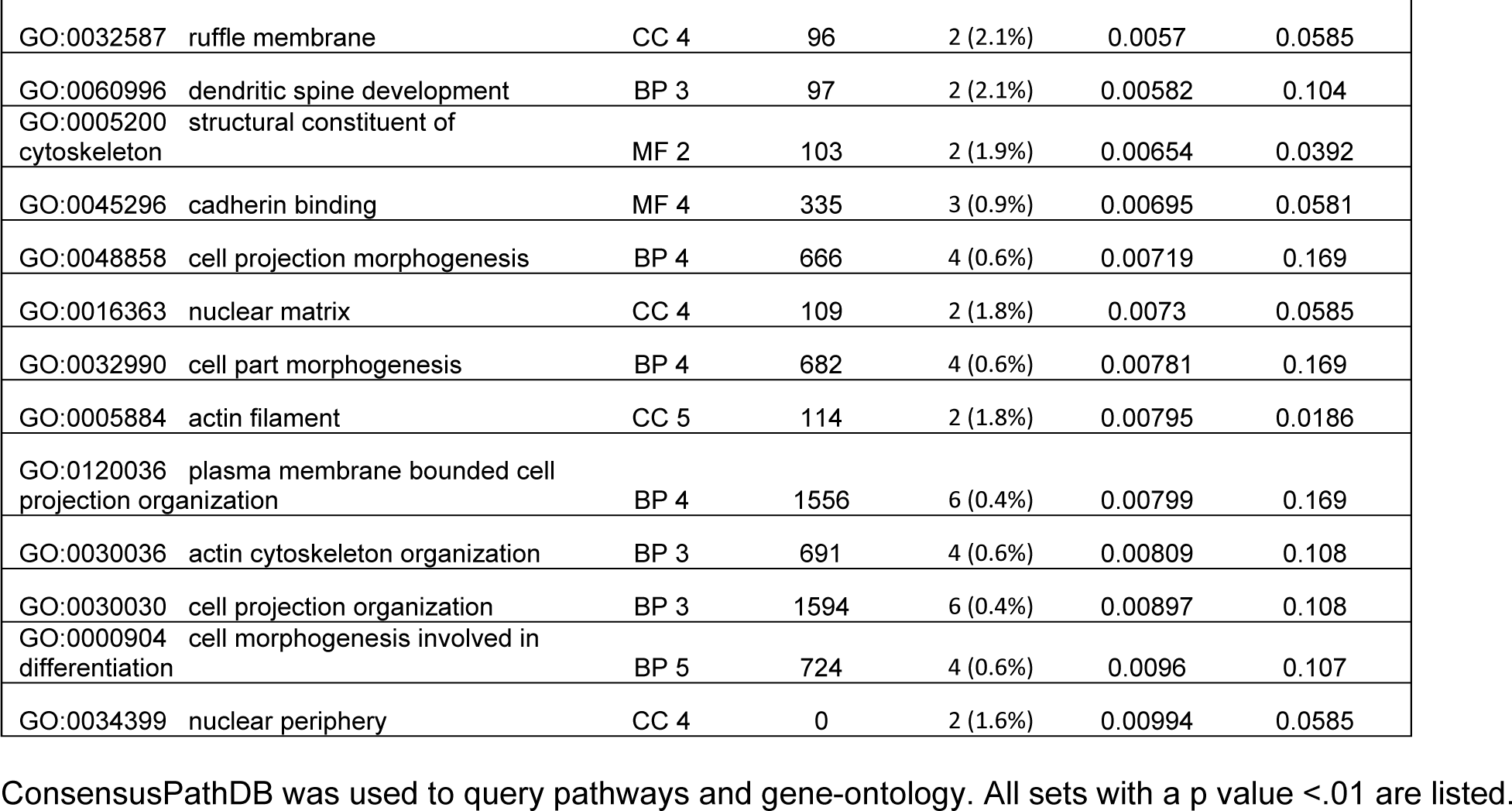
Genes harboring ultra-rare de novo variants in ADHD cases are enriched for pathway and gene ontology based sets

| Enriched pathway-based sets |  |  |  |  |  |
| --- | --- | --- | --- | --- | --- |
| pathway name | set size | candidates contained | p-value | q-value | pathway source |
| CXCR4-mediated signaling events | 86 | 3 (3.6%) | 0.000152 | 0.00414 | PID |
| Sema3A PAK dependent Axon repulsion | 16 | 2 (12.5%) | 0.00018 | 0.00414 | Reactome |
| Ectoderm Differentiation | 142 | 3 (2.1%) | 0.00071 | 0.00856 | Wikipathways |
| EPHB-mediated forward signaling | 35 | 2 (5.7%) | 0.000879 | 0.00856 | Reactome |
| rac1 cell motility signaling pathway | 36 | 2 (5.6%) | 0.00093 | 0.00856 | BioCarta |
| RAC1 signaling pathway | 54 | 2 (3.7%) | 0.00208 | 0.016 | PID |
| Semaphorin interactions | 64 | 2 (3.1%) | 0.00292 | 0.0192 | Reactome |
| CDC42 signaling events | 71 | 2 (2.8%) | 0.00357 | 0.0198 | PID |
| EPH-Ephrin signaling | 74 | 2 (2.7%) | 0.00388 | 0.0198 | Reactome |
| Fc gamma R-mediated phagocytosis - Homo sapiens (human) | 97 | 2 (2.1%) | 0.00644 | 0.0296 | KEGG |
| Enriched gene ontology-based sets |  |  |  |  |  |
| gene ontology term | category, level | set size | candidates contained | p-value | q-value |
| GO:0044877 protein-containing complex binding | MF 2 | 1223 | 7 (0.6%) | 0.000384 | 0.00461 |
| GO:0000226 microtubule cytoskeleton organization | BP 3 | 615 | 5 (0.8%) | 0.000653 | 0.0559 |
| GO:0005856 cytoskeleton | CC 4 | 2323 | 9 (0.4%) | 0.000816 | 0.022 |
| GO:0007010 cytoskeleton organization | BP 4 | 1405 | 7 (0.5%) | 0.000879 | 0.0993 |
| GO:0040019 positive regulation of embryonic development | BP 5 | 42 | 2 (4.8%) | 0.00112 | 0.0564 |
| GO:0050808 synapse organization | BP 3 | 410 | 4 (1.0%) | 0.00126 | 0.0559 |
| GO:0050807 regulation of synapse organization | BP 5 | 210 | 3 (1.4%) | 0.00188 | 0.0564 |
| GO:0050803 regulation of synapse structure or activity | BP 3 | 220 | 3 (1.4%) | 0.00214 | 0.0574 |
| GO:0060997 dendritic spine morphogenesis | BP 5 | 59 | 2 (3.4%) | 0.0022 | 0.0564 |
| GO:0007017 microtubule-based process | BP 2 | 808 | 5 (0.6%) | 0.00221 | 0.112 |
| GO:0016358 dendrite development | BP 3 | 235 | 3 (1.3%) | 0.00258 | 0.0574 |
| GO:0015629 actin cytoskeleton | CC 5 | 513 | 4 (0.8%) | 0.00281 | 0.0112 |
| GO:0015630 microtubule cytoskeleton | CC 5 | 1292 | 6 (0.5%) | 0.00321 | 0.0112 |
| GO:0044430 cytoskeletal part | CC 3 | 1777 | 7 (0.4%) | 0.00342 | 0.0478 |
| GO:0005912 adherens junction | CC 3 | 557 | 4 (0.7%) | 0.00382 | 0.0478 |
| GO:0097061 dendritic spine organization | BP 4 | 81 | 2 (2.5%) | 0.0041 | 0.169 |
| GO:0070161 anchoring junction | CC 2 | 573 | 4 (0.7%) | 0.00423 | 0.114 |
| GO:0031334 positive regulation of protein complex assembly | BP 5 | 287 | 3 (1.0%) | 0.00453 | 0.0775 |
| GO:0008013 beta-catenin binding | MF 3 | 86 | 2 (2.3%) | 0.00461 | 0.0737 |
| GO:0106027 neuron projection organization | BP 5 | 90 | 2 (2.2%) | 0.00503 | 0.0775 |

|  |  |  |  |  |  |  |
| --- | --- | --- | --- | --- | --- | --- |
| GO:0032587 | ruffle membrane | CC 4 | 96 | 2 (2.1%) | 0.0057 | 0.0585 |
| GO:0060996 | dendritic spine development | BP 3 | 97 | 2 (2.1%) | 0.00582 | 0.104 |
| GO:0005200 | structural constituent of cytoskeleton | MF 2 | 103 | 2 (1.9%) | 0.00654 | 0.0392 |
| GO:0045296 | cadherin binding | MF 4 | 335 | 3 (0.9%) | 0.00695 | 0.0581 |
| GO:0048858 | cell projection morphogenesis | BP 4 | 666 | 4 (0.6%) | 0.00719 | 0.169 |
| GO:0016363 | nuclear matrix | CC 4 | 109 | 2 (1.8%) | 0.0073 | 0.0585 |
| GO:0032990 | cell part morphogenesis | BP 4 | 682 | 4 (0.6%) | 0.00781 | 0.169 |
| GO:0005884 | actin filament | CC 5 | 114 | 2 (1.8%) | 0.00795 | 0.0186 |
| GO:0120036 | plasma membrane bounded cell projection organization | BP 4 | 1556 | 6 (0.4%) | 0.00799 | 0.169 |
| GO:0030036 | actin cytoskeleton organization | BP 3 | 691 | 4 (0.6%) | 0.00809 | 0.108 |
| GO:0030030 | cell projection organization | BP 3 | 1594 | 6 (0.4%) | 0.00897 | 0.108 |
| GO:0000904 | cell morphogenesis involved in differentiation | BP 5 | 724 | 4 (0.6%) | 0.0096 | 0.107 |
| GO:0034399 | nuclear periphery | CC 4 | 0 | 2 (1.6%) | 0.00994 | 0.0585 |

ConsensusPathDB was used to query pathways and gene-ontology. All sets with a p value <.01 are listed.

**Dataset S1 (separate file).** Metrics and quality control information from whole-exome DNA sequencing data. Key included in dataset.

**Dataset S2 (separate file).** Details of rare *de novo* variants identified in ADHD probands and controls. Key included in dataset.

**Dataset S3 (separate file).** Gene-based test results combining the ultra-rare *de novo* damaging variants and independent case-control data. We used the Bayesian extension of the Transmission And De novo Association test (extTADA) to examine ultra-rare de novo protein-truncating variants (PTV) and missense variants predicted to be damaging (MPC score>2, Mis-D) from the 147 ADHD parent-child trios and an independent group of 3,206 ADHD cases and 5,002 unaffected controls. We ran extTADA to calculate the Bayes factor and q-values (false discovery rate, FDR) for each gene. One gene *KDM5B* is classified as a high-confidence risk gene (FDR, false discovery rate < 0.1) and two genes, *POMT1* and *YLPM1*, are classified as probable risk genes (FDR<0.3).

## References

1. G. V. Polanczyk, G. A. Salum, L. S. Sugaya, A. Caye, L. A. Rohde, Annual research review: A meta-analysis of the worldwide prevalence of mental disorders in children and adolescents. Journal of child psychology and psychiatry 56, 345–365 (2015).

2. J. Posner, G. V. Polanczyk, E. Sonuga-Barke, Attention-deficit hyperactivity disorder. Lancet 395, 450–462 (2020).

3. S. V. Faraone, H. Larsson, Genetics of attention deficit hyperactivity disorder. Molecular psychiatry 24, 562–575 (2019).

4. D. Demontis et al., Genome-wide analyses of ADHD identify 27 risk loci, refine the genetic architecture, and implicate several cognitive domains. Nature genetics, 1–11 (2023).

5. D. Demontis et al., Discovery of the first genome-wide significant risk loci for attention deficit/hyperactivity disorder. Nature genetics 51, 63–75 (2019).

6. E. J. Sonuga-Barke et al., Annual Research Review: Perspectives on progress in ADHD science–from characterization to cause. Journal of Child Psychology and Psychiatry (2022).

7. B. Harich et al., From rare copy number variants to biological processes in ADHD. American Journal of Psychiatry 177, 855–866 (2020).

8. F. K. Satterstrom et al., Autism spectrum disorder and attention deficit hyperactivity disorder have a similar burden of rare protein-truncating variants. Nat Neurosci 22, 1961–1965 (2019).

9. F. K. Satterstrom et al., Large-scale exome sequencing study implicates both developmental and functional changes in the neurobiology of autism. Cell 180, 568–584 e523 (2020).

10. J. Kaplanis et al., Evidence for 28 genetic disorders discovered by combining healthcare and research data. Nature 586, 757–762 (2020).

11. S. Wang et al., De novo sequence and copy number variants are strongly associated with tourette disorder and implicate cell polarity in pathogenesis. Cell reports 24, 3441–3454 e3412 (2018).

12. A. C. Lionel et al., Rare copy number variation discovery and cross-disorder comparisons identify risk genes for ADHD. Science translational medicine 3, 95ra75-95ra75 (2011).

13. J. Martin et al., A brief report: de novo copy number variants in children with attention deficit hyperactivity disorder. Transl Psychiatry 10, 135 (2020).

14. B. R. Al-Mubarak et al., Whole exome sequencing in ADHD trios from single and multi-incident families implicates new candidate genes and highlights polygenic transmission. European Journal of Human Genetics 28, 1098–1110 (2020).

15. L. de Araújo Lima et al., An integrative approach to investigate the respective roles of single-nucleotide variants and copy-number variants in Attention-Deficit/Hyperactivity Disorder. Scientific reports 6, 22851 (2016).

16. D. S. Kim et al., Sequencing of sporadic Attention-Deficit Hyperactivity Disorder (ADHD) identifies novel and potentially pathogenic de novo variants and excludes overlap with genes associated with autism spectrum disorder. American Journal of Medical Genetics Part B: Neuropsychiatric Genetics 174, 381–389 (2017).

17. M. Halvorsen et al., Exome sequencing in obsessive-compulsive disorder reveals a burden of rare damaging coding variants. Nat Neurosci 24, 1071–1076 (2021).

18. E. Olfson et al., Whole-exome DNA sequencing in childhood anxiety disorders identifies rare de novo damaging coding variants. Depress Anxiety 39, 474–484 (2022).

19. K. E. Samocha et al., Regional missense constraint improves variant deleteriousness prediction bioRxiv 10.1101/148353 (2017).

20. H. T. Nguyen et al., Integrated Bayesian analysis of rare exonic variants to identify risk genes for schizophrenia and neurodevelopmental disorders. Genome medicine 9, 1–22 (2017).

21. G. Zhao et al., Gene4Denovo: an integrated database and analytic platform for de novo mutations in humans. Nucleic acids research 48, D913–D926 (2020).

22. J. Harrington, G. Wheway, S. Willaime-Morawek, J. Gibson, Z. S. Walters, Pathogenic KDM5B variants in the context of developmental disorders. Biochimica et Biophysica Acta (BBA)-Gene Regulatory Mechanisms, 194848 (2022).

23. V. Faundes et al., Histone lysine methylases and demethylases in the landscape of human developmental disorders. The American Journal of Human Genetics 102, 175–187 (2018).

24. H. C. Martin et al., Quantifying the contribution of recessive coding variation to developmental disorders. Science (New York, N.Y.) 362, 1161–1164 (2018).

25. T. Geis et al., Clinical long-time course, novel mutations and genotype-phenotype correlation in a cohort of 27 families with POMT1-related disorders. Orphanet Journal of Rare Diseases 14, 1–17 (2019).

26. D. Song et al., Genetic variations and clinical spectrum of dystroglycanopathy in a large cohort of Chinese patients. Clinical genetics 99, 384–395 (2021).

27. M. Medina, R. C. Marinescu, J. Overhauser, K. S. Kosik, Hemizygosity of δ-catenin (CTNND2) is associated with severe mental retardation in cri-du-chat syndrome. Genomics 63, 157–164 (2000).

28. T. N. Turner et al., Loss of δ-catenin function in severe autism. Nature 520, 51–56 (2015).

29. A.-F. van Rootselaar et al., δ-Catenin (CTNND2) missense mutation in familial cortical myoclonic tremor and epilepsy. Neurology 89, 2341–2350 (2017).

30. A. Gregor et al., De novo variants in the F-box protein FBXO11 in 20 individuals with a variable neurodevelopmental disorder. The American Journal of Human Genetics 103, 305–316 (2018).

31. S. Jansen et al., De novo variants in FBXO11 cause a syndromic form of intellectual disability with behavioral problems and dysmorphisms. European Journal of Human Genetics 27, 738–746 (2019).

32. D. Lehalle et al., STAG1 mutations cause a novel cohesinopathy characterised by unspecific syndromic intellectual disability. Journal of medical genetics 54, 479–488 (2017).

33. R. Karlsson Linnér et al., Multivariate analysis of 1.5 million people identifies genetic associations with traits related to self-regulation and addiction. Nature neuroscience 24, 1367–1376 (2021).

34. V. Trubetskoy et al., Mapping genomic loci implicates genes and synaptic biology in schizophrenia. Nature 604, 502–508 (2022).

35. C. Cappi et al., De novo damaging DNA coding mutations are associated with obsessive-compulsive disorder and overlap with Tourette’s disorder and autism. Biological psychiatry 87, 1035–1044 (2020).

36. G. D. Fischbach, C. Lord, The Simons Simplex Collection: a resource for identification of autism genetic risk factors. Neuron 68, 192–195 (2010).

37. W. J. Chen, S. V. Faraone, J. Biederman, M. T. Tsuang, Diagnostic accuracy of the Child Behavior Checklist scales for attention-deficit hyperactivity disorder: a receiver-operating characteristic analysis. Journal of consulting and clinical psychology 62, 1017 (1994).

38. A. McKenna et al., The Genome Analysis Toolkit: a MapReduce framework for analyzing next-generation DNA sequencing data. Genome research 20, 1297–1303 (2010).

39. K. Wang, M. Li, H. Hakonarson, ANNOVAR: functional annotation of genetic variants from high-throughput sequencing data. Nucleic acids research 38, e164 (2010).

40. A. Manichaikul et al., Robust relationship inference in genome-wide association studies. Bioinformatics (Oxford, England) 26, 2867–2873 (2010).

41. K. J. Karczewski et al., The mutational constraint spectrum quantified from variation in 141,456 humans. Nature 581, 434–443 (2020).

42. P. Feliciano et al., Exome sequencing of 457 autism families recruited online provides evidence for autism risk genes. NPJ Genom Med 4, 19 (2019).

43. A. Buniello et al., The NHGRI-EBI GWAS Catalog of published genome-wide association studies, targeted arrays and summary statistics 2019. Nucleic acids research 47, D1005–D1012 (2019).

44. E. Sollis et al., The NHGRI-EBI GWAS Catalog: knowledgebase and deposition resource. Nucleic acids research 51, D977–D985 (2023).

45. R. Herwig, C. Hardt, M. Lienhard, A. Kamburov, Analyzing and interpreting genome data at the network level with ConsensusPathDB. Nature protocols 11, 1889–1907 (2016).

## SI References

1. H. T. Nguyen et al., Integrated Bayesian analysis of rare exonic variants to identify risk genes for schizophrenia and neurodevelopmental disorders. Genome medicine 9, 1-22 (2017).

2. X. He et al., Integrated model of de novo and inherited genetic variants yields greater power to identify risk genes. PLoS genetics 9, e1003671 (2013).

3. J. S. Ware, K. E. Samocha, J. Homsy, M. J. Daly, Interpreting de novo Variation in Human Disease Using denovolyzeR. Curr Protoc Hum Genet 87, 7.25.21-27.25.15 (2015).

